# Design and implementation of maternal-infant clinical trial recruitment alert using linked electronic medical records, and evaluation of researcher-perceived alert usability

**DOI:** 10.64898/2026.06.30.26356791

**Authors:** Haustine Patt Panganiban, Ahuva Segal, Carl Kuschel

## Abstract

**Background:** Clinical decision support systems configured as electronic medical records alerts can support clinical trial recruitment by identifying eligible participants. While they are typically used to evaluate a single patient record, their application for a linked maternal-infant chart has not been extensively explored.

**Objective:** This project aimed to design and implement a clinical trial alert using linked maternal–infant records and to evaluate researchers’ perceived usability.

**Materials and Methods:** We conducted a two-phase quality assurance project: (1) design and implementation of an alert aligned with the anticipated recruitment workflow, and (2) evaluation of usability using the System Usability Scale. Basic content analysis described the alert design and implementation processes, while quantitative scoring assessed perceived usability.

**Results:** Over a 12-month period, only one alert was triggered due to changes in the recruitment workflow. Two silent alerts assessed maternal eligibility in outpatient and infant eligibility in inpatient settings. Three of four researchers completed the survey, yielding a score of 92.5, indicating excellent usability.

**Conclusion:** Although the alert was technically functional and perceived to have excellent usability, its performance was limited by deviations from the intended recruitment workflow. Researcher engagement and recruitment workflow alignment emerged as critical factors influencing alert utility. Clinical trial alerts for linked maternal–infant records can be designed and implemented with excellent usability; however, consistent adherence to the recruitment workflow is essential. Broader application in additional study settings is recommended.

## Background

The widespread adoption of electronic medical records (EMR) has created new opportunities to improve the conduct of clinical trials[1,2]. By providing a digital clinical infrastructure, EMR enable the integration of tools such as clinical decision support systems (CDSS), which can generate automated alerts and streamline clinical trial recruitment workflows [2–6]. These systems have been employed in multiple studies to facilitate clinical trial recruitment by evaluating eligibility using routinely collected clinical data from medical records [3–5,7–12].

However, the application of CDSS to linked patient records, particularly linked maternal and infant records, remains underexplored. Within the EMR, maternal and infant records exist as separate entities: the maternal record is created antenatally, while the infant record is created at birth and subsequently linked to the maternal record. Screening for clinical trial eligibility across these linked records typically requires manual review of both maternal and infant records, often spanning multiple encounters, making the process time-consuming and labour-intensive [13,14]. This needs to review two distinct, yet clinically related entities, continues to present a barrier to efficient clinical trial recruitment.

We conducted a quality assurance project to address gaps in clinical trial alerts for linked mother-infant records. We aimed to design and implement an alert using the linked maternal-infant EMR and to assess researchers’ perceptions of the alert’s usability. We anticipated that our findings could inform future initiatives and support the implementation of projects with similar requirements within our organization and beyond. This project was reviewed and approved by The Royal Women’s Research Office (reference number QA/115362/RWHV-2025-464833).

We conducted this project in the Newborn Research Centre at The Royal Women’s Hospital (The Women’s), a state-wide tertiary hospital in Melbourne, Australia. We utilised the EMR (Epic EHR Systems, Verona, USA), which was implemented organization-wide in August 2020. The Royal Women’s Hospital is part of the Parkville Electronic Medical Record precinct, which operates a single installation across The Royal Melbourne Hospital, Peter MacCallum Cancer Centre and The Royal Children’s Hospital.

## Methods

We used a combination of basic content analysis to describe the design and implementation process, and basic quantitative analysis to assess the researchers’ perceived alert usability for this project. We followed the STARE-HI (Statement on Reporting of Evaluation Studies in Health Informatics) guidelines for reporting the project findings [15].

## Study Design

We focused on an EMR alert developed to support the “BLUEPRINT: Predicting Long-term Outcomes of Prematurity from Early Life Events” (ACTRN12624000851561) study timeline (Figure 1). This study was selected as it required a linked maternal-infant EMR to be evaluated for participant eligibility.

**Figure 1.**
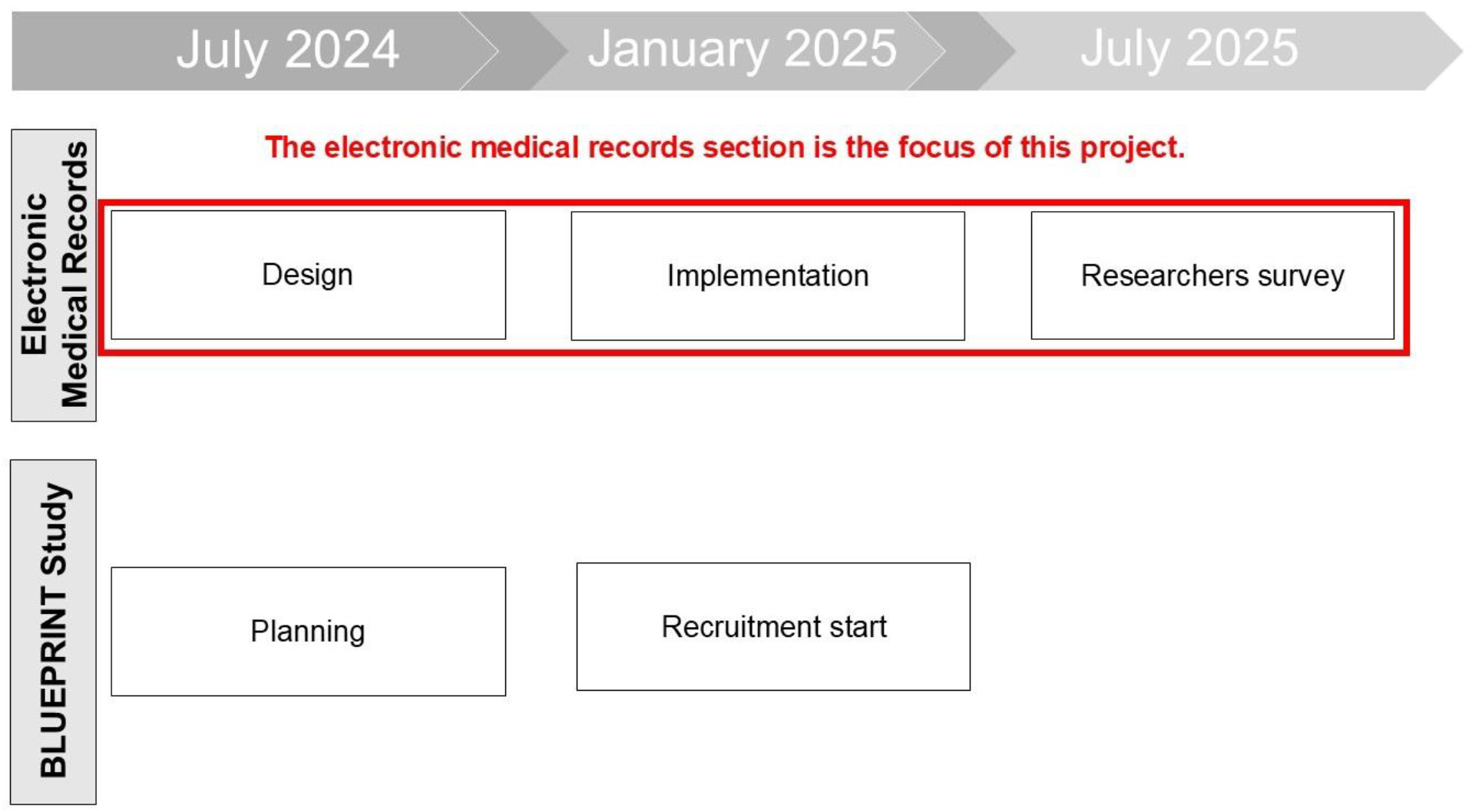
BLUEPRINT: Predicting Long-term Outcomes of Prematurity from Early Life Events” study timeline

We divided this project into two phases. The first phase described the design and implementation of the alert and was conducted from March 1, 2025, to July 31, 2025. We collected data from the EMR used to build the alert and conducted a basic content analysis.

The second phase involved sending an online survey using the System Usability Scale (SUS) [16] questionnaire to evaluate the researcher’s perceived alert usability. The survey contained 10 standard SUS questions with minimal modifications (Supplementary Material). The modified SUS is a ten-item Likert scale that assesses system usability, with each options assigned a value (strongly disagree = 1, disagree = 2, neutral = 3, agree = 4 and strongly agree = 5) [16]. The word “system” was changed to “alert” to be consistent with the CDSS terminology used for this project. This conducted from July 1, 2025, to August 31, 2025, and used REDCap [17] to create and distribute the survey. The BLUEPRINT study team, composed of investigators and coordinators, were sent an invitation with a participant information sheet. The collected anonymous data were calculated using the formula: SUS score = (X-5 + 25-Y) x 2.5, where X is the sum of the points for all odd-numbered questions and Y is the sum of the points for all even-numbered questions, which yields a SUS acceptability score out of 100 [16]. The curved grading scale for SUS [18] was used to analyze the SUS score, which provides a total score from 0 (poor usability) to 100 (excellent usability).

## RESULTS

### Design process

The alert design was undertaken by a multidisciplinary team through a co-design approach, incorporating hospital clinicians, researchers, and EMR analysts with complementary expertise spanning clinical trial methodology and EMR build and implementation.

We created two silent alerts for this project based on the study recruitment workflow (Figure 2). In this workflow, the study coordinator conducts an eligibility assessment of the mother’s record prior to birth. If the mother meets the eligibility criteria, the record is associated with the study. Following delivery, the clinician creates the infant’s record, which is automatically linked to the maternal record. Once the infant’s record is created, the predefined alert criteria are systematically evaluated. If the record satisfies all criteria, the EMR automatically sends a message and notification to the study team.

**Figure 2.**
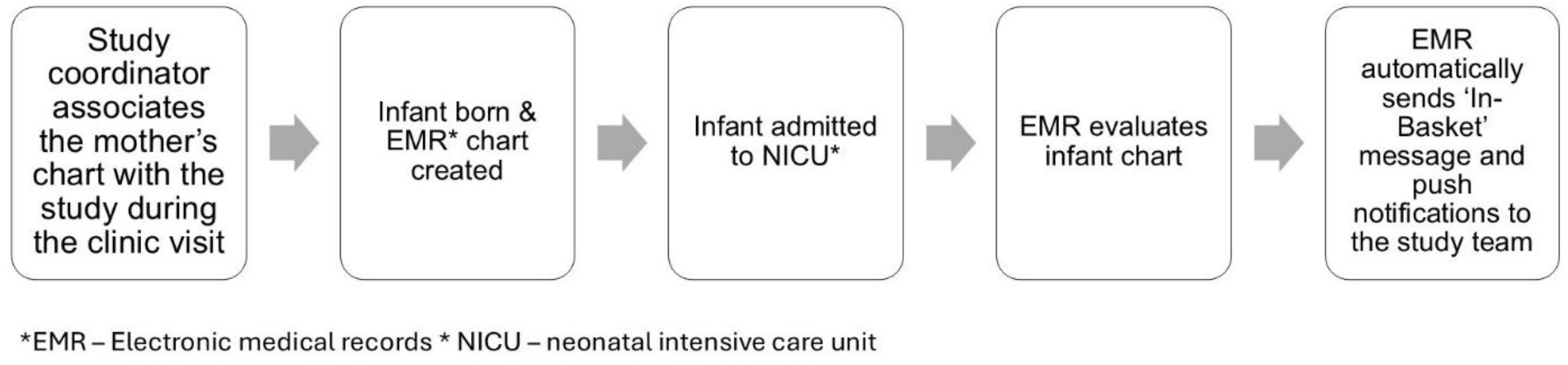
Study recruitment workflow

The first silent alert (Figure 3) incorporated a criterion that assesses the maternal record to verify its linkage to a study record with confirmed consent status. The study record is a feature within the EMR that allows the study team to monitor the participant’s progress through associating the participant with the study. The silent alert was triggered when a user opened the maternal record. Although the alert was triggering silently, we set a 9999-hour lockout with a maximum of 1 pop-up during the lockout to prevent a system performance issue.

**Figure 3.**
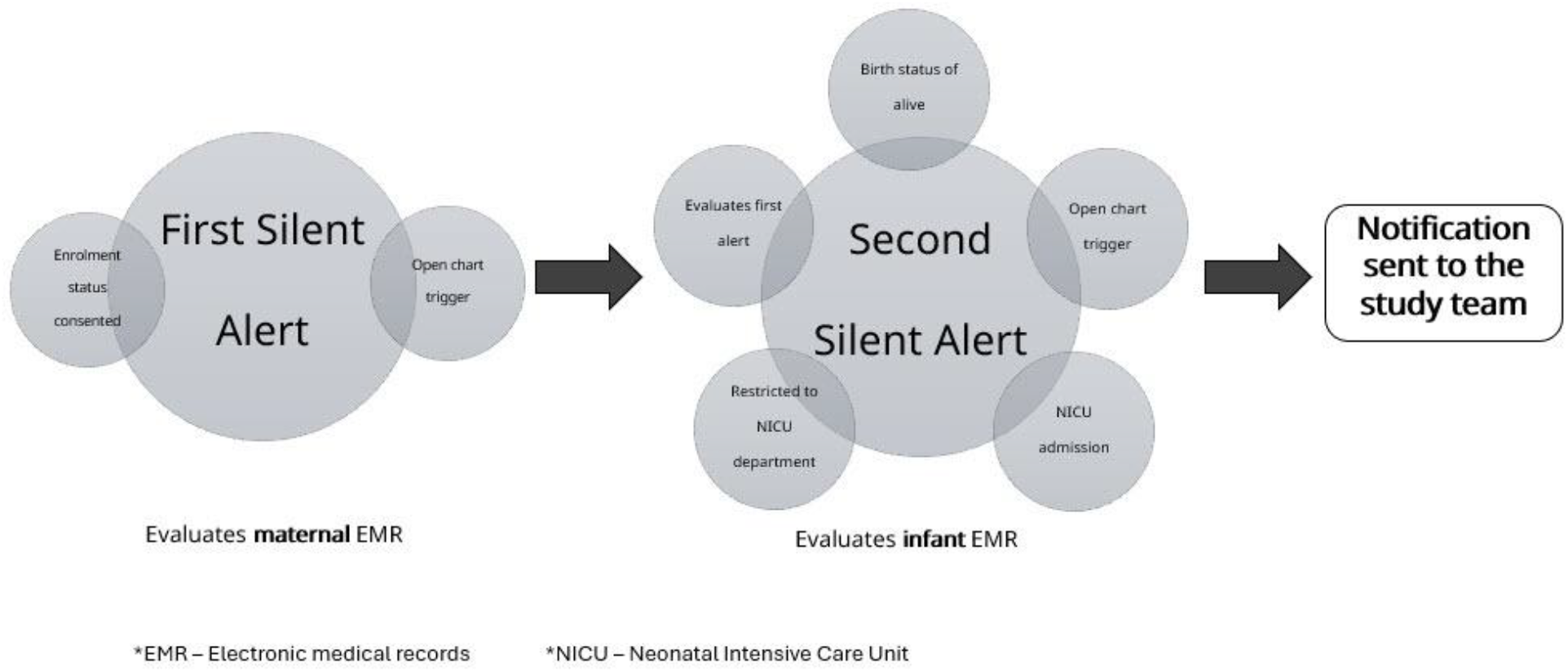
Alert design

The second silent alert (Figure 3) evaluated the infant record to determine whether the linked mother record was associated with the study record. Other criteria checked whether the infant was currently admitted to the neonatal intensive care unit (NICU), and the patient was not deceased.

We configured the alert to send a message to the study team using the email-like messaging functionality within the EMR and a dedicated folder was created, and notification subject lines were standardized. The infant’s demographic and clinical information was also displayed to the study team to assist in screening and monitoring (Supplementary Material).

We designed the alert according to the principles of right person, right format, and right workflow, consistent with commonly accepted methodologies for clinical decision support [19]. The proof-of-concept design prioritized user experience and followed an iterative development process [20]. The final proof-of-concept was co-designed and iteratively refined in collaboration with the study team, who approved the final design.

### Implementation process

The alert was approved by local governance procedures, including review at both the local health service and the broader precinct. Study team training was delivered primarily via e-mail communication, with additional ad hoc training provided through in-person and online sessions as required. We deployed the alert within the EMR on December 11, 2024, after which post-implementation monitoring was undertaken to identify incidents and issues. We scheduled follow-up with the study team that occurred weekly during the first month post-implementation, and monthly for the subsequent six months. The alert behaviour was audited using standard EMR reporting tools.

### Researcher-Perceived Alert Usability

We assessed perceived usability by a survey sent to 4 study members, comprising 2 investigators and 2 study coordinators, on July 7, 2025. Three responses were received. The overall alert usability scale grade was 92.5, which corresponds ‘excellent usability’ based on the curved grading scale for the SUS [18].

From December 11, 2024, to December 11, 2025, 49 participants were enrolled in the study. However, only one EMR alert was triggered and sent to the study team via the anticipated recruitment workflow. Following consultation with the study team, it became apparent that the recruitment was primarily occurring during infant admission to the NICU, rather than during the prenatal visits for which the alert has been configured. The difference between the assumed and actual recruitment workflow substantially reduced the anticipated alert rate. The project lead undertook a revalidation of the alert design, which demonstrated that the alert was working as designed based on the recruitment workflow in Figure 3 and no modifications to the existing design were necessary. To encourage adherence to the workflow, information sessions were delivered to the study team to reinforce the purpose and advantages of the alert system.

## DISCUSSION

In this quality assurance project, we described the design and implementation process for an alert using a linked maternal–infant EMR and evaluated researchers’ perceived alert usability. Our project yielded two key findings that may help address the barrier to participant recruitment posed by maternal and infant eligibility screening. First, a clinical trial alert can leverage the relationship between linked maternal–infant records, which is distinctive from alerts previously reported in the existing literature and may serve as a foundation for future implementation. Second, the findings from researchers’ perceptions of clinical trial alert usability contribute to the limited literature in this area.

In this project, study team engagement played a pivotal role in designing a meaningful alert tailored to the anticipated recruitment workflow. Similar to the team composition described by Austrian et al. [3], we collaborated with the study team and precinct partners to enhance alert efficiency. Although the literature increasingly supports the use of alerts for clinical and research workflows, there are also reports of negative outcomes, such as alert fatigue [7,12] and system noise [3]. To mitigate these issues, we designed the alert to be non-interruptive with an appropriate trigger, similar to that described by Melnick et al. [21], ensuring support for the clinical trial workflow without introducing unnecessary noise. Additionally, we implemented a lockout time to suppress repeated alert triggers, preventing system performance issues and reducing alert volume - an underlying challenge in designing effective CDSS [3]. To enhance effectiveness of the EMR, we also configured the alert to send a real-time message that served as a notification to prompt the study team to review the infant’s record [6].

In terms of implementation, our project followed a series of structured processes. Our organization is part of a precinct comprising four hospitals sharing EMR infrastructure, where optimization changes require change control approval. This process was smooth and ensured that the project adhered to established change management procedures and guidelines. Training and communication for this project were seamless, as only a limited number of users were affected. We provided training both online and through elbow-to-elbow support, which facilitated the smooth implementation of the alert. We established ongoing feedback sessions with the study team during implementation, similar to the approach described by Austrian et al. [3] and Deakyne et al. [6], which helped tailor the alert to suit the recruitment workflow. The feedback session also provided an opportunity to address issues encountered during implementation and maintained study team engagement to ensure compliance with the study protocol.

Although only one alert was sent to the study team, our findings indicate that the alert itself was perceived as useful and demonstrated excellent usability. However, this finding cannot be benchmarked against current literature [22] due to differences in population, low number of alerts triggered, and system design, highlighting the need for future studies to explore researchers’ perceptions of alert usability. Our analysis did not examine the relationship between alert frequency and recruitment rate due to the change in recruitment strategy, which deviated from the intended workflow, resulting in a low number of alerts firing. Despite the study team involvement in the alert design, reinforcing the alert’s purpose and maintaining continuous engagement with the study team, no increase in the alert firing rate was observed.

Our project has several limitations. This project was conducted in a single centre and single site, using a single study model. This limitation also extends to the selection of survey participants, which was restricted to members of the study team. As a next step, we plan to apply the alert design to additional research studies to determine whether it can be implemented across multiple studies and to assess the alert’s perceived usability among other study teams.

## Data Availability

All data produced in the study are available upon reasonable request to the authors.

## CONCLUSION

Clinical trial alerts can be designed and implemented using linked maternal–infant EMR, with researchers reporting excellent perceived usability. Despite these positive outcomes, deviations from the intended recruitment workflow resulted in minimal alert firing and limited assessment of its overall impact. Future work should validate this alert design and implementation approach across multiple studies and study teams to further evaluate usability and broader applicability.

## AUTHOR CONTRIBUTIONS

HP conceptualized and led this project. AS and CK contributed to the project protocol development. HP conducted the data collection and analysis. HP drafted the manuscript and AS and CK critically revised the manuscript.

## COMPETING INTEREST

None declared.

## DECLARATION OF GENERATIVE AI and AI-ASSISTED TECHNOLOGIES

During the preparation of this manuscript, the author(s) used Microsoft Copilot to assist with minor language editing. Following use of this tool, the author(s) critically reviewed and revised the content as necessary and assume full responsibility for the final published version

## LEGENDS

**Supplementary Material 1A.**
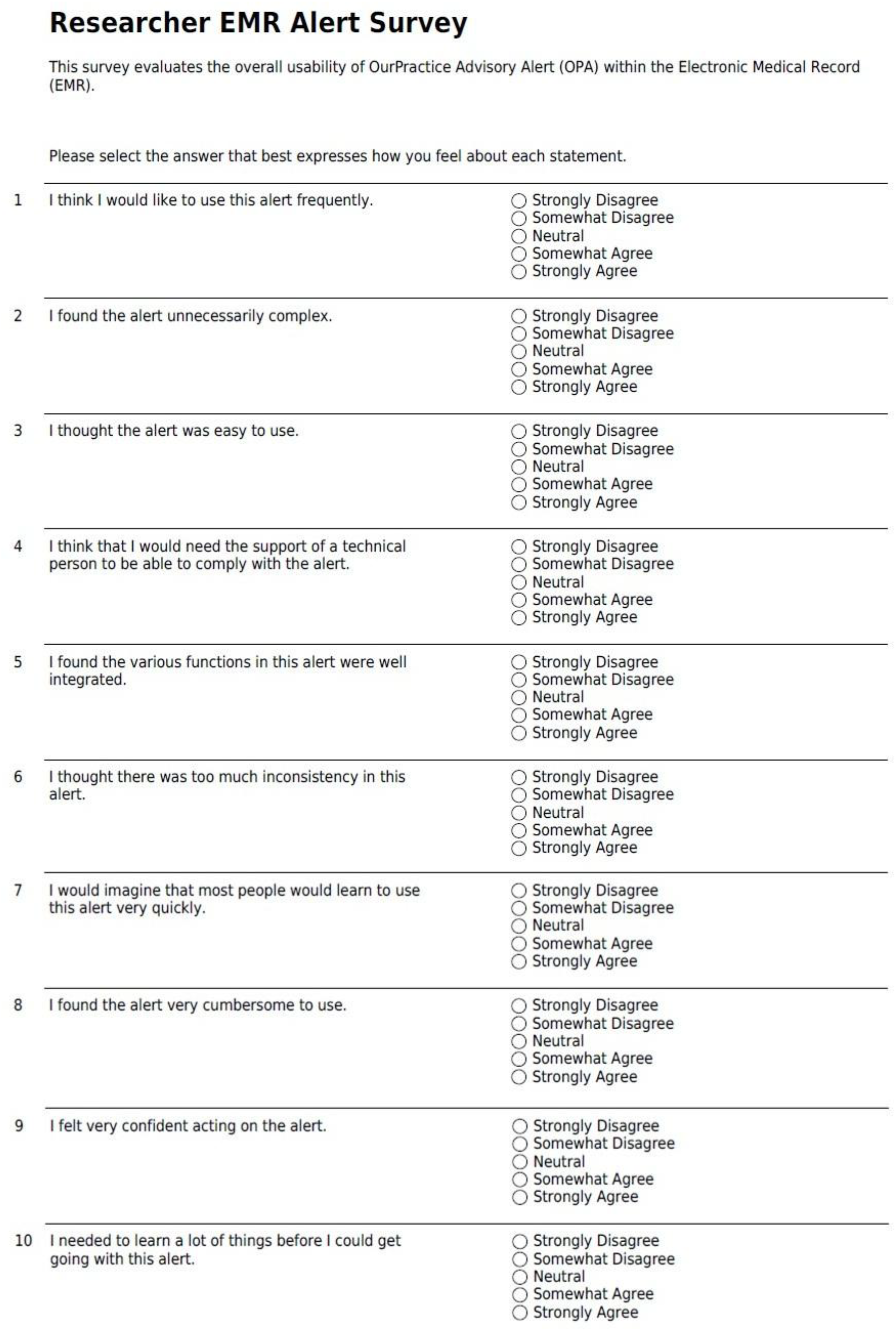
Modified system usability scale (SUS) for researchers

**Supplementary Material 1B.**
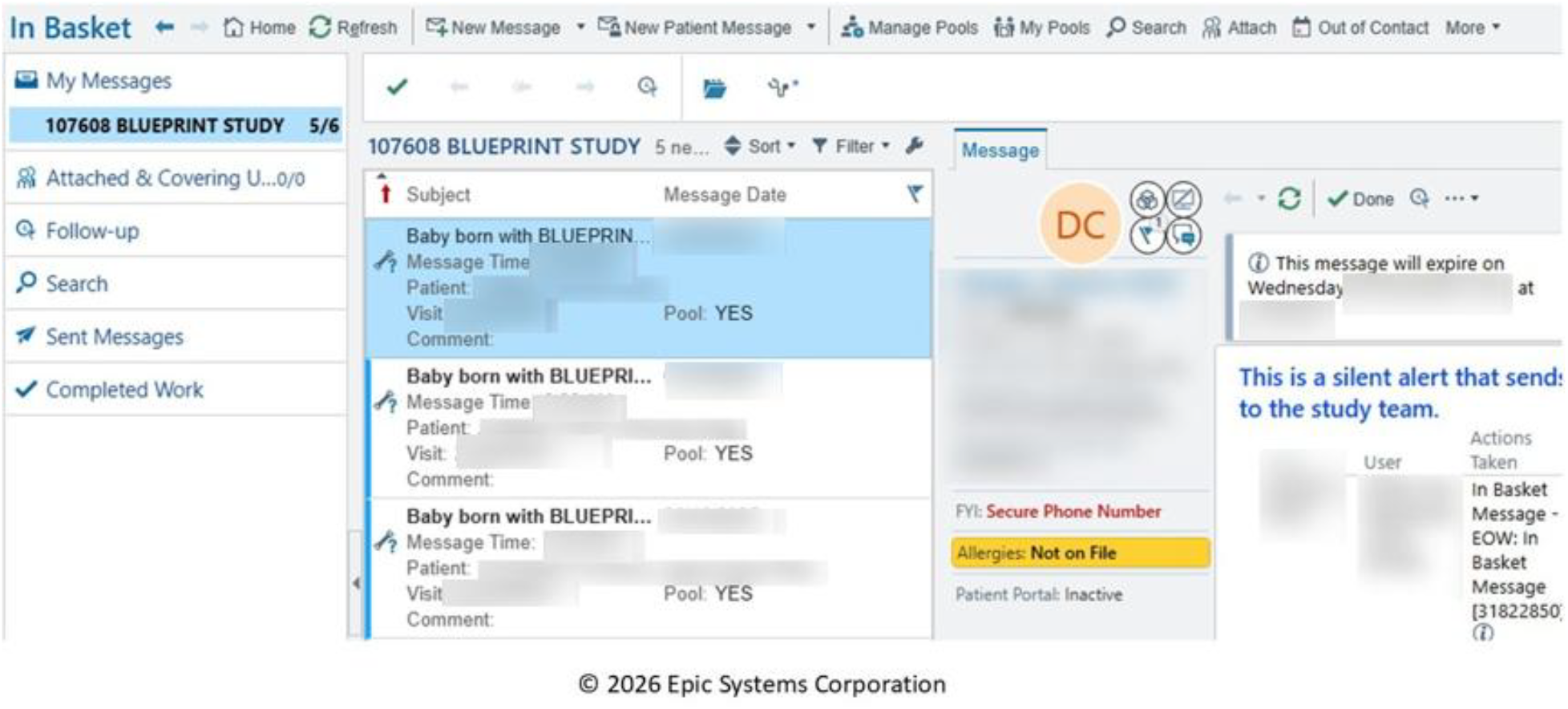
Message sent to the study team

## Notes

### Competing Interest Statement

The authors have declared no competing interest.

### Clinical Trial

ACTRN12624000851561

### Author Declarations

Ethics committee of The Royal Womens Hospital gave ethical approval for this work.

