## Supplementary Material for "Design and implementation of maternal-infant clinical trial recruitment alert using linked electronic medical records, and evaluation of researcher-perceived alert usability"

**Supplementary Material 1 A** Modified system usability scale (SUS) for researchers

### Researcher EMR Alert Survey

This survey evaluates the overall usability of OurPractice Advisory Alert (OPA) within the Electronic Medical Record (EMR).

Please select the answer that best expresses how you feel about each statement.

| | | |
|---|---|---|
| 1 | I think I would like to use this alert frequently. | ◯ Strongly Disagree<br>◯ Somewhat Disagree<br>◯ Neutral<br>◯ Somewhat Agree<br>◯ Strongly Agree |
| 2 | I found the alert unnecessarily complex. | ◯ Strongly Disagree<br>◯ Somewhat Disagree<br>◯ Neutral<br>◯ Somewhat Agree<br>◯ Strongly Agree |
| 3 | I thought the alert was easy to use. | ◯ Strongly Disagree<br>◯ Somewhat Disagree<br>◯ Neutral<br>◯ Somewhat Agree<br>◯ Strongly Agree |
| 4 | I think that I would need the support of a technical person to be able to comply with the alert. | ◯ Strongly Disagree<br>◯ Somewhat Disagree<br>◯ Neutral<br>◯ Somewhat Agree<br>◯ Strongly Agree |
| 5 | I found the various functions in this alert were well integrated. | ◯ Strongly Disagree<br>◯ Somewhat Disagree<br>◯ Neutral<br>◯ Somewhat Agree<br>◯ Strongly Agree |
| 6 | I thought there was too much inconsistency in this alert. | ◯ Strongly Disagree<br>◯ Somewhat Disagree<br>◯ Neutral<br>◯ Somewhat Agree<br>◯ Strongly Agree |
| 7 | I would imagine that most people would learn to use this alert very quickly. | ◯ Strongly Disagree<br>◯ Somewhat Disagree<br>◯ Neutral<br>◯ Somewhat Agree<br>◯ Strongly Agree |
| 8 | I found the alert very cumbersome to use. | ◯ Strongly Disagree<br>◯ Somewhat Disagree<br>◯ Neutral<br>◯ Somewhat Agree<br>◯ Strongly Agree |
| 9 | I felt very confident acting on the alert. | ◯ Strongly Disagree<br>◯ Somewhat Disagree<br>◯ Neutral<br>◯ Somewhat Agree<br>◯ Strongly Agree |
| 10 | I needed to learn a lot of things before I could get going with this alert. | ◯ Strongly Disagree<br>◯ Somewhat Disagree<br>◯ Neutral<br>◯ Somewhat Agree<br>◯ Strongly Agree |

**Supplementary Material 1 B** Message sent to the study team

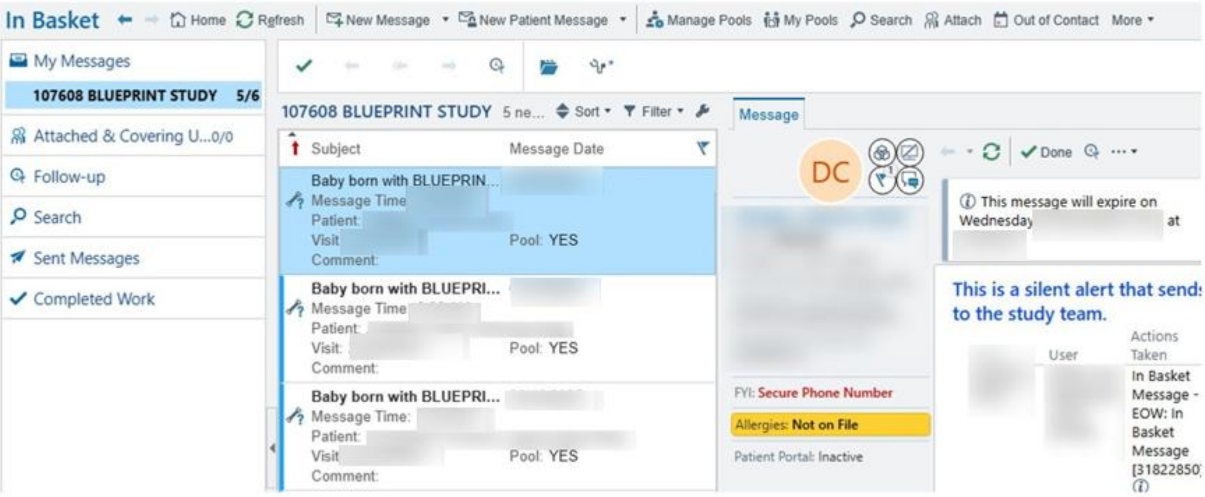


© 2026 Epic Systems Corporation
